# Colocalization of expression transcripts with COVID-19 outcomes is rare across cell states, cell types and organs

**DOI:** 10.1101/2023.05.29.23290694

**Authors:** Julian Daniel Sunday Willett, Tianyuan Lu, Tomoko Nakanishi, Satoshi Yoshiji, Guillaume Butler-Laporte, Sirui Zhou, Yossi Farjoun, J. Brent Richards

**Affiliations:** Centre for Clinical Epidemiology, Department of Medicine, Lady Davis Institute, Jewish General Hospital, McGill University, Montréal, Québec, Canada; McGill University, Montreal, Quebec, Canada; Quantitative Life Sciences Program, McGill University, Montreal, Quebec, Canada; Department of Human Genetics, McGill University, Montreal, Quebec, Canada; Kyoto-McGill International Collaborative Program in Genomic Medicine, Graduate School of Medicine, Kyoto University, Kyoto, Japan; Research Fellow, Japan Society for the Promotion of Science, Tokyo, Japan; Genome Centre, McGill University, Montreal, Quebec, Canada; Departments of Medicine, Human Genetics, Epidemiology and Biostatistics, McGill University, Montréal, Quebec, Canada; Department of Twin Research, King’s College London, London, United Kingdom; Five Prime Sciences Inc, Montréal, Québec, Canada

**Author notes:** Corresponding author: Brent Richards, Pavillon H-413, Jewish General Hospital 3755 Cote Ste Catherine, Montreal, QC, Canada, H3T 1E2.

**Keywords:** COVID-19, colocalization, Mendelian randomization, eQTL, scRNA

## Abstract

Identifying causal genes at GWAS loci can help pinpoint targets for therapeutic interventions. Expression studies can disentangle such loci but signals from expression quantitative trait loci (eQTLs) often fail to colocalize—which means that the genetic control of measured expression is not shared with the genetic control of disease risk. This may be because gene expression is measured in the wrong cell type, physiological state, or organ. We tested whether Mendelian randomization (MR) could identify genes at loci influencing COVID-19 outcomes and whether the colocalization of genetic control of expression and COVID-19 outcomes was influenced by cell type, cell stimulation, and organ.

We conducted MR of *cis*-eQTLs from single cell (scRNA-seq) and bulk RNA sequencing. We then tested variables that could influence colocalization, including cell type, cell stimulation, RNA sequencing modality, organ, symptoms of COVID-19, and SARS-CoV-2 status among individuals with symptoms of COVID-19. The outcomes used to test colocalization were COVID-19 severity and susceptibility as assessed in the Host Genetics Initiative release 7.

Most transcripts identified using MR did not colocalize when tested across cell types, cell state and in different organs. Most that did colocalize likely represented false positives due to linkage disequilibrium. In general, colocalization was highly variable and at times inconsistent for the same transcript across cell type, cell stimulation and organ. While we identified factors that influenced colocalization for select transcripts, identifying 33 that mediate COVID-19 outcomes, our study suggests that colocalization of expression with COVID-19 outcomes is partially due to noisy signals even after following quality control and sensitivity testing. These findings illustrate the present difficulty of linking expression transcripts to disease outcomes and the need for skepticism when observing eQTL MR results, even accounting for cell types, stimulation state and different organs.

**Author Summary:** The genetic determinants of disease and gene expression often do not colocalize (which means they do not share a single causal signal). While some researchers have identified factors that could explain this disconnect, such as immune stimulation or tissue studied, understanding of this complex phenomenon remains incomplete. A deeper understanding could help identify additional genes that mediate disease, affording promising targets for treatment or prevention of disease. We used RNA sequencing data collected at the single cell and bulk tissue level to identify genes whose expression influenced COVID-19 outcomes. We assessed which variables influencing colocalization, including cell type, cell stimulation, RNA sequencing modality, organ, symptoms of COVID-19, and SARS-CoV-2 status among individuals with symptoms of COVID-19. We observed that colocalization of specific candidate genes identified by MR was highly variable and influenced by multiple factors, including cell state and cell population. These results illustrate that even after assessing multiple variables that may influence colocalization, there existed few examples of genes identified by MR that colocalized with gene expression. Future studies would benefit from larger transcriptomics study cohorts and more advanced statistical methods which better account for differences in linkage disequilibrium panels between data sources.

## Introduction

Severe COVID-19 is partially influenced by immune hyperstimulation (Merad et al. 2022; Tan et al. 2021). The immune response is mediated by different immune cell subtypes, acting at varying time points during infection across tissues (Ong et al. 2020; Tan et al. 2021). Understanding the dynamics of this process may pinpoint targets helpful for COVID-19 interventions, as previously shown (De Biasi et al. 2020; Kundu et al. 2022; Mathew et al. 2020).

One way to investigate mechanisms influencing COVID-19 outcomes is to determine the underlying contributory genetic factors. GWAS has identified 87 loci associated with COVID-19 outcomes, but it is often unclear which gene(s) at such loci drive this association (Covid-19 Host Genetics Initiative 2021). Resolving a GWAS locus to its causal gene(s) is non-trivial (Forgetta et al. 2022). One way to identify causal genes at GWAS loci is to examine whether associated SNPs influence outcomes in an appropriate cell type. However, genetic determinants of gene expression (expression quantitative trait loci; eQTL) have often failed to “colocalize” with disease outcomes (Connally et al. 2022). Colocalization, in this context, means that gene expression and the disease outcome share a single common causal SNP (Connally et al. 2022). This lack of colocalization is concerning and not fully resolved. However, given the central importance of gene expression in disease incidence and progression, efforts are required to explain the paradox that the genetic determinants of gene expression often appear different than the those of disease, even for known causal genes in known causal cell types or tissues (Connally et al. 2022). We sought to determine if this lack of colocalization could be explained when cell type, cell stimulation, method of sequencing and organ were taken into account.

One factor that may influence colocalization is the population of cells studied. Gene expression is typically determined in bulk tissue, which provides a mixture of cells from the tissue. Such signal dilution, combined with complex factors such as cell-cell interactions, may explain why bulk tissue eQTLs often fail to colocalize with disease outcomes (Connally et al. 2022). Single-cell sequencing studies (scRNA-seq) assay gene expression in specific cell types and thus single-cell sequencing can provide a less heterogeneous assessment of gene expression. Comparing colocalization between bulk and single-cell sequencing, studying additional variables that influence the association, could resolve the contributory factors. This knowledge could help identify causal genes at GWAS loci and accelerate drug development by targeting pathways causal for disease. Targets supported by MR and colocalization evidence are more likely to anticipate clinical trial results, where the target of the medicine in the trial is a circulating protein and its causal influence upon disease is supported by both MR and colocalization. (Zheng et al. 2020).

Several studies have investigated the relationship between gene expression and COVID-19 outcomes using older releases of expression data or COVID-19 outcomes. Pairo-Castineira *et al*. found that increased *TYK2* and decreased *IFNAR2* expression in whole blood were associated with life-threatening COVID-19 (Pairo-Castineira et al. 2021). Schmiedel *et al*. found several genes whose expression in specific immune cell types and tissues, including resting and activated naive CD4+ cells, influenced and colocalized with genetic determinants of COVID- 19 outcomes (Schmiedel et al. 2021). D’Antonio et al. found genes that colocalized with COVID-19 loci in whole blood, including *ABO* and *IFNAR2*, and identified the causal variants using fine-mapping (D’Antonio et al. 2021).

Recently, Soskic *et al*. profiled the changes in gene expression in CD4+ T-cells following stimulation with anti-CD3/anti-CD28 human T-activators (Soskic et al. 2022). We aimed to determine if cell and cell-state specific gene expression could identify novel determinants of COVID-19 outcomes, suggesting which cells are responsible for COVID-19 mortality risk and when. Further, we aimed to determine if such cellular specificity may clarify colocalization of expression and GWAS data. Repeating this analysis in other cell types, bulk whole-blood in individuals with symptoms of COVID-19, with and without a recent PCR-confirmed infection, and bulk tissues in individuals assessed prior to the pandemic lacking any symptoms would identify differences in colocalization due to sequencing modality and different clinical states. The results would afford insights into whether the genetic control of gene expression and disease risk is clarified when resolving to single cells, specific cellular states and clinical characteristics of patients sampled.

To answer these questions, we undertook a four-stage study design (**Fig. 1**). First, we conducted Mendelian randomization (MR) of *cis*-eQTLs obtained from single-cell RNA-sequencing (scRNA-seq) data from CD4+ T-cell subtypes at varying times after stimulation with gene expression as exposures and COVID-19 severity as an outcome. Second, we tested these MR-identified genes for colocalization with COVID-19 outcomes across all time points following stimulation of CD4+ T-cells. Third, we compared these colocalization results to those from bulk whole-blood RNA sequencing obtained from individuals with COVID-19 symptoms, who were either SARS-CoV-2 PCR positive or negative. Fourth, we compared the colocalization results to bulk unstimulated whole blood and 47 other tissues obtained from individuals assessed in GTEx v.8, whose tissues were apparently undiseased at time of sampling.

**Fig. 1.**
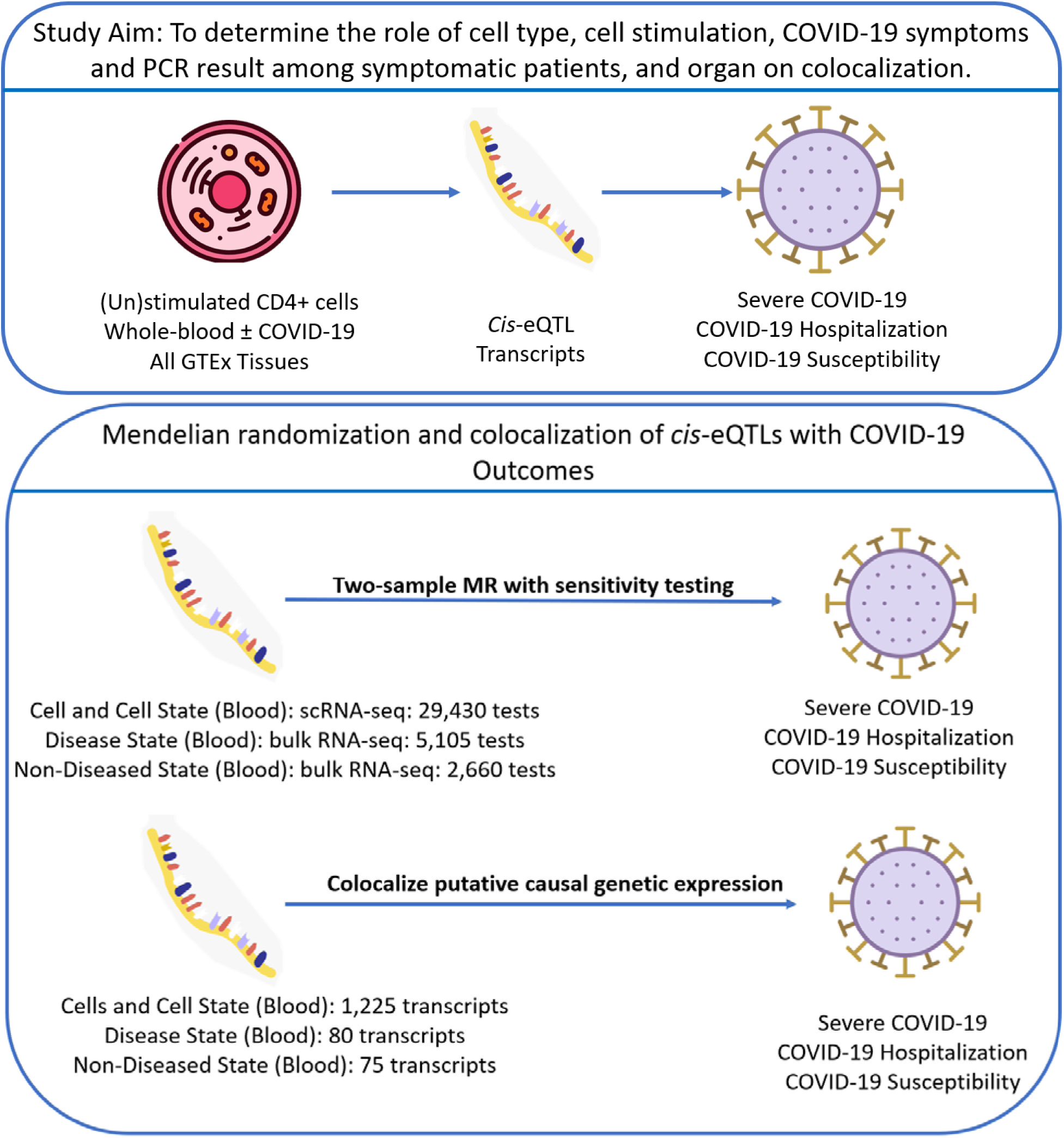
Study overview

These findings identified 33 genes whose expression may influence COVID-19 outcomes. Colocalization was highly variable and not consistent across the factors that may influence it. These results underline the complexity of factors that influence the colocalization of genetic expression with disease.

## Results

### Cohort demographics

All datasets used in this study included individuals solely of European ancestry to reduce potential confounding by population stratification. ScRNA-seq data for CD4+ T cells in whole blood was obtained from Soskic *et al*., who isolated cells from 119 healthy, British-ancestry individuals, with a mean age of 47 years, where 44% were females (Soskic et al. 2022). Bulk whole-blood RNA sequencing of individuals with symptoms of COVID-19, who were SARS-CoV-2 positive or negative, was obtained from BQC19, a Quebec cohort of individuals recruited from hospitals presenting with COVID-19 symptoms. BQC19 RNA-sequencing data comprised 112 individuals with symptoms of COVID-19 and SARS-CoV-2 positive PCR tests and 166 individuals who also had symptoms of COVID-19 but a SARS-CoV-2 negative PCR test. The mean age of BQC19 participants was 54 years and 53% were females. The GTEx Consortium cohort comprised 838 post-mortem donors, including 715 individuals of European ancestry, of which 33.5% were female (GTEx Consortium 2020).

### MR and sensitivity testing

To identify genes influencing COVID-19 outcomes, we used either a Wald ratio or inverse-variance weighted MR analysis at 81 non-MHC COVID-19 associated loci (**Supplemental Table 1**) across three COVID-19 outcomes (severe disease, hospitalized disease, and susceptibility to disease) with sensitivity analyses for each test, detailed in **Methods**. We identified 33 genes whose expression was shown by MR to influence COVID-19 severity and susceptibility (**Fig. 2**, **Table 1, Supplemental Table 2**).

**Fig. 2.**
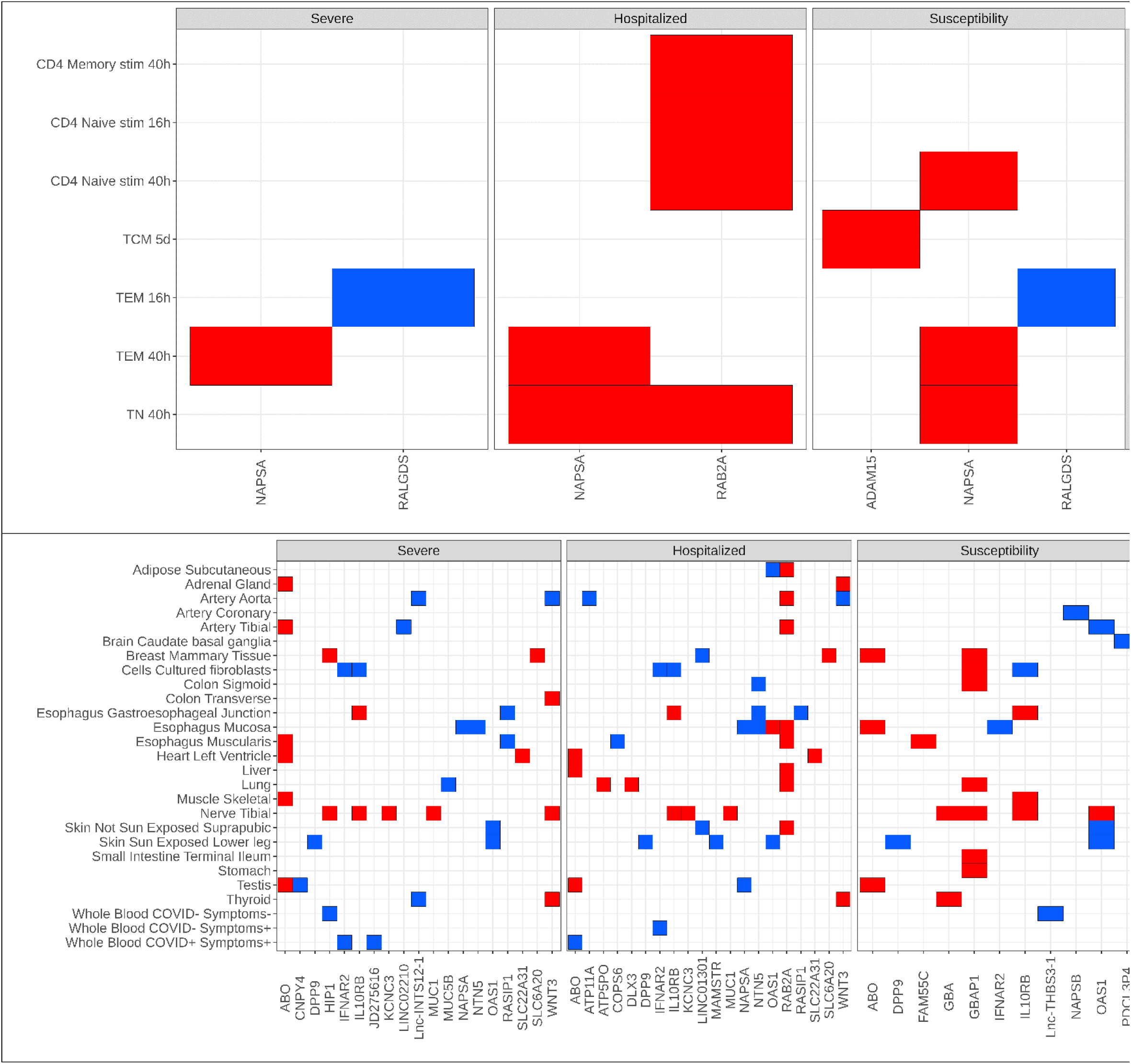
A total of 33 genes that colocalized across body tissue, evaluated at the (a) single-cell and (b) bulk tissue level, were estimated to have their expression increase (red) and decrease (blue) risk of COVID-19 outcomes. In bulk tissue, COVID+ referred to individuals who had tested positive for COVID-19 with COVID- for those who tested negative and Symptoms+ refers to individuals who presented with symptoms of COVID-19 with Symptoms- referring to those who did not have symptoms of COVID-19. Note that some transcripts show increased expression in some tissues to be associated with COVID-19 outcomes, whereas other tissues show decreased expression to be associated with the same transcript for the same outcome. TCM: T central memory cell. TEM: T effector memory cell. TN: T naïve cell.

**Table 1.**
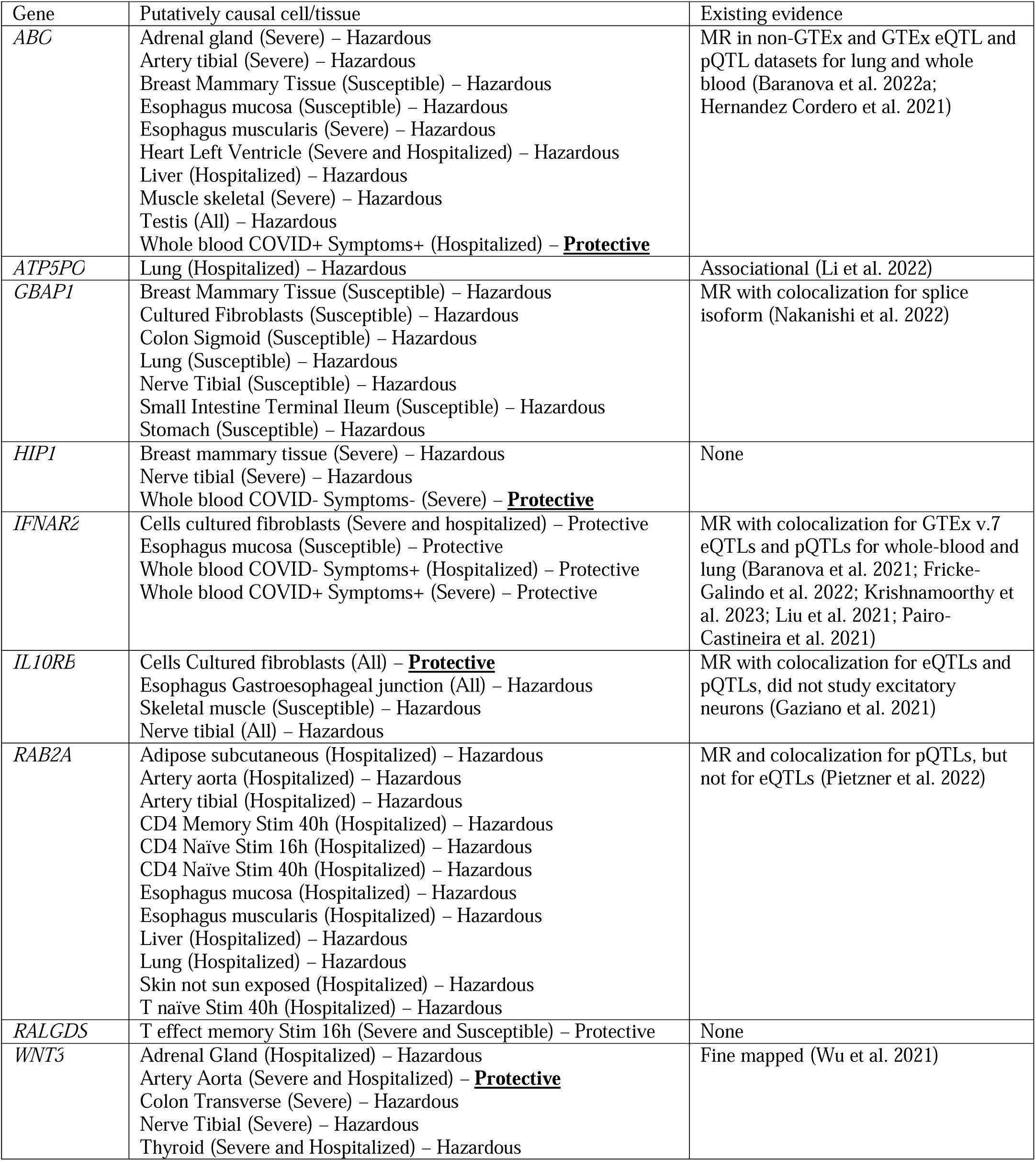
Putatively causal associations of gene expression with COVID-19 outcomes. While some genes had opposite effects on risk for a minority of tissues (bolded), the direction of effect of most was consistent when observed in multiple tissues.

### Single-cell results

Of the 29,430 combinations of CD4+ cell:stimulation-state:gene:outcome for the whole-blood single-cell eQTLs, 1,225 transcripts (4.2%) had a multiple-testing, Benjamini-Hochberg adjusted p-value ≤ 0.05 in the MR analyses. We limited results to only those arising from *cis*-eQTLs to reduce potential bias from horizontal pleiotropy. *Cis*-eQTLs were defined as genetic variants associated with transcript level within ±500 kb of the transcriptional start site. We also only retained *cis*-eQTLs that were within ±500 kb of the lead SNP associated with a COVID-19 outcomes (**Supplemental Table 3)**. Only 13 of these 1,225 *cis*-eQTLs passed colocalization sensitivity testing, defined as having a probability of the gene’s expression and the COVID-19 outcome sharing a single causal variant (PP_H4_) greater than 0.80 (**Fig. 1-2**). These data demonstrate that only a small proportion of *cis*-single-cell eQTLs identified via MR colocalized with COVID-19 outcomes, suggesting that such MR findings do not consistently reflect a common causal genetic signal shared between the transcript and COVID-19 outcomes.

### Bulk results

Next, we assessed *cis*-eQTLs from the bulk whole-blood RNA sequencing in the BQC19 cohort, which comprised individuals with symptoms of COVID-19 who had either a positive, or negative PCR test for SARS-CoV-2. Of the 5,105 combinations of study-group:gene:outcome for the BQC19 *cis*-eQTLs, 80 transcripts (1.6%) were identified by MR to have effects on COVID-19 outcomes (**Supplemental Table 4)**. Only 4 of these 80 transcripts passed colocalization sensitivity testing (**Fig. 1-2**). We next assessed whether we would observe similar results if we used whole blood and other organ bulk RNA sequencing from GTEx v.8 (GTEx Consortium 2020) as the source of the cis-eQTLs, that included individuals without symptoms or seropositivity for COVID-19. In GTEx bulk whole-blood, we found that of the 2,660 combinations of gene:outcome for the GTEx *cis*-eQTLs, 75 (2.8%) were identified by MR testing to have effects upon COVID-19 outcomes, with only two colocalizing (**Fig. 1-2**). In all 48 GTEx tissues, we found that of the 125,088 combinations of tissue:gene:outcome tested, 3190 (2.6%) were estimated to influence COVID-19 outcomes by MR but only 98 of these colocalized (**Fig. 2**).

Thus, taken together, across the three different sources of gene expression data (scRNA-seq whole blood CD4+ T-cells, bulk whole blood RNA sequencing in patients with symptoms of COVID-19, and bulk RNA sequencing in individuals whose tissues were apparently health across 48 tissues, including whole blood), we observed 33 unique putatively causal transcripts across 115 specific states that colocalized, which represent 2.3% of those transcripts that survived MR testing and multiple testing thresholds. These findings suggest that most transcripts identified to be associated with COVID-19 outcomes via MR fail to colocalize even across single cell and bulk sequencing, as well as different cellular states and patient states.

### MR implicated several cis-eQTLs that increase or decrease the risk for COVID-19 outcomes with few overlapping transcripts between scRNA-seq and bulk RNA sequencing results

Across outcomes, all cell types from the scRNA-seq whole-blood data had at least one *cis*-eQTL estimated to be causal for a COVID-19 outcome via MR, except T effector memory re-expressing CD45RA and T regulatory cells (**Fig. 2**). Of the four genes estimated to have colocalized causal effects from the whole-blood scRNA-seq MR experiments, *RALGDS* expression was the only *cis*- eQTL that decreased the risk of COVID-19 outcomes. Specifically, *RALGDS* expression reduced the risk of severe disease (OR = 0.78, 95% CI: 0.71-0.87; adjusted p = 1.1 x 10^-4^) and susceptibility to disease (OR = 0.88, 95% CI: 0.86-0.90; adjusted p = 7.4 x 10^-18^), when expressed in solely T effector memory cells 16 hours after stimulation (**Fig. 2**). *RALGDS’* influence on hospitalized COVID-19 was not clearly different from the null (OR = 0.87, 95% CI: 0.79-0.96; adjusted p = 0.08) but passed MR sensitivity testing and colocalized. *NAPSA* expression in T naive cells 40 hours after stimulation increased risk of hospitalized (OR = 1.17, 95% CI: 1.08-1.28; adjusted p = 5.0 x 10^-3^) and susceptibility to (OR = 1.08, 95% CI: 1.05-1.11; adjusted p = 1.7 x 10^-5^) COVID-19 (**Fig. 2**). *NAPSA’*s influence on severe COVID-19 was not different from the null (OR = 1.20, 95% CI: 1.06-1.37; adjusted p = 0.07) but passed MR sensitivity testing and colocalized. *NAPSA* also increased risk for every outcome in T effector memory cells 40 hours after stimulation (OR = 1.18, 95% CI: 1.10-1.27; adjusted p = 4.0 x 10^-4^ for severe, OR = 1.15, 95% CI: 1.10-1.20; adjusted p = 5.9 x 10^-9^ for hospitalized, OR = 1.04, 95% CI: 1.03-1.06; adjusted p = 1.2 x 10^-4^ for susceptibility) (**Fig. 2**).

Interestingly, none of the colocalizing transcripts from scRNA-seq overlapped with colocalizing bulk RNA sequencing *cis*-eQTLs from BQC19 or GTEx in matching tissues. Generally, signals in bulk sequencing were harder to separate from noise, compared to single-cell results. Increased *IFNAR2* expression was protective in individuals with symptoms of COVID-19 without PCR-confirmed SARS-CoV-2 against severe COVID-19, or individuals who were negative for or had perhaps not yet tested positive for COVID-19 (OR = 0.75, 95% CI: 0.66-0.86; adjusted p = 3.0 x 10^-3^) with insufficient evidence to suggest protection for individuals who did test positive (adjusted p = 0.04) where it failed weighted mode MR sensitivity testing but colocalized. In contrast, *IFNAR2* was protective for only individuals who tested positive for SARS-CoV-2 against hospitalized COVID-19 (OR = 0.86, 95% CI: 0.80-0.94; adjusted p = 0.03) with insufficient evidence to suggest protection for BQC19 SARS-CoV-2 negative individuals (adjusted p = 8.1 x 10^-3^) where it failed MR Egger intercept sensitivity testing (p = 0.02) while colocalizing (**Fig. 2**). This was perhaps a false negative. There were several other genes that colocalized for only a subset of outcomes in other tissues, although some consistently colocalized in the same tissue across outcomes, such as ABO in the testis and DPP9 in sun-exposed skin (**Table 1, Supplemental Table 2**).

### Colocalization of specific MR-identified cis-eQTLs depended on cell stimulation

To investigate the variables that influenced colocalization, we conducted colocalization on each gene that was estimated causal in MR. Most MR-identified transcripts did not colocalize (**Fig. 1, Supplemental Tables 6-8)**. The proportion of estimated causal transcripts that colocalized and passed single causal variant sensitivity testing was 1.1% for whole blood single-cell eQTLs, 5.0% for BQC19, 2.7% for GTEx whole blood, and 3.1% for all organs in GTEx (**Fig. 3**). Colocalization of 2/4 single-cell eQTLs was specific to cell type (**Fig. 2**) with 3/3 from stimulated cells specific to cell state with colocalization for *RALGDS*, for example, specific to T effector memory cells 16 hours post-stimulation for severe and susceptibility to COVID-19 (**Fig. 4**).

**Fig 3.**
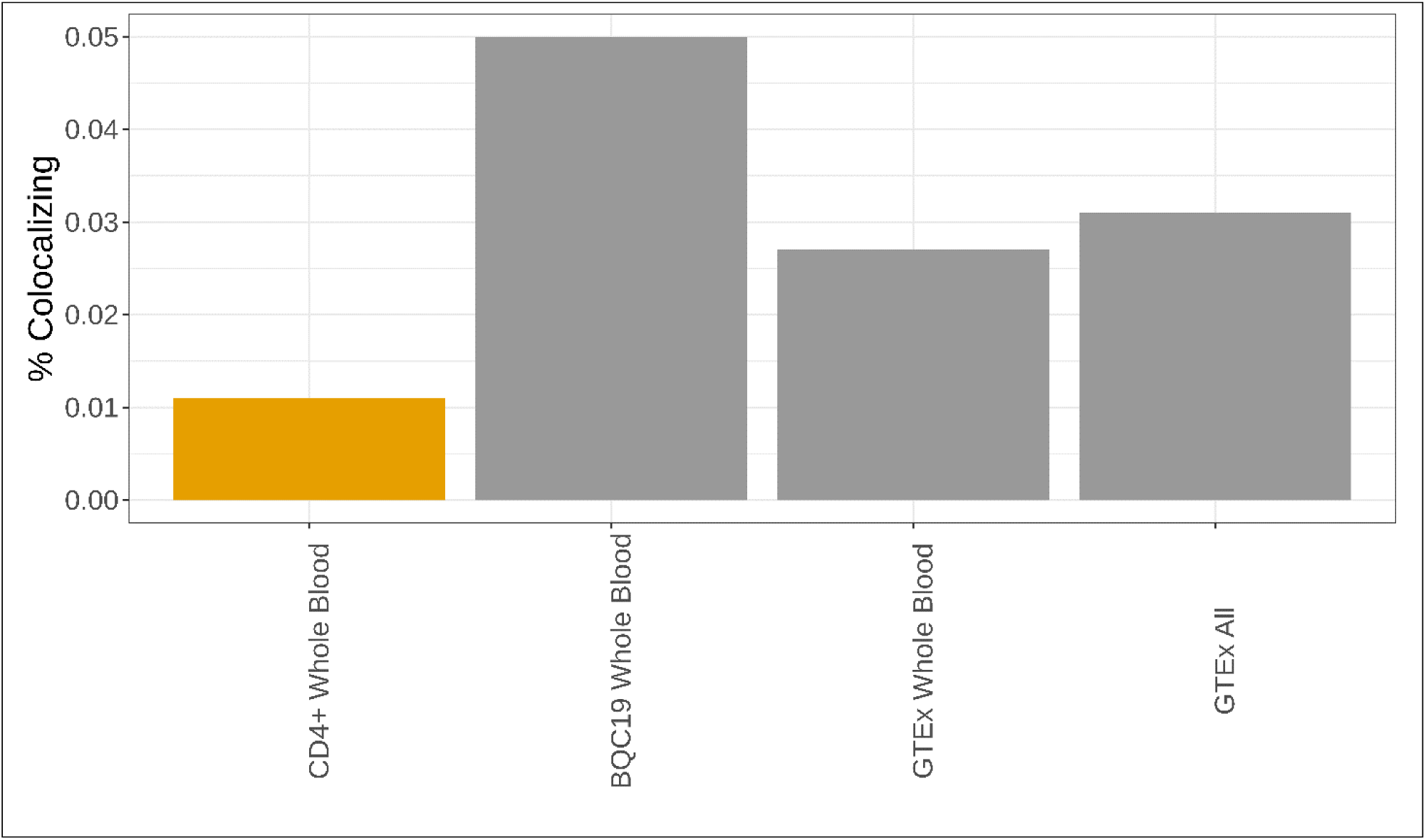
The proportion of estimated causal variants that colocalized and passed sensitivity testing. Orange bars refer to single cell data, gray bulk sequencing.

**Fig 4.**
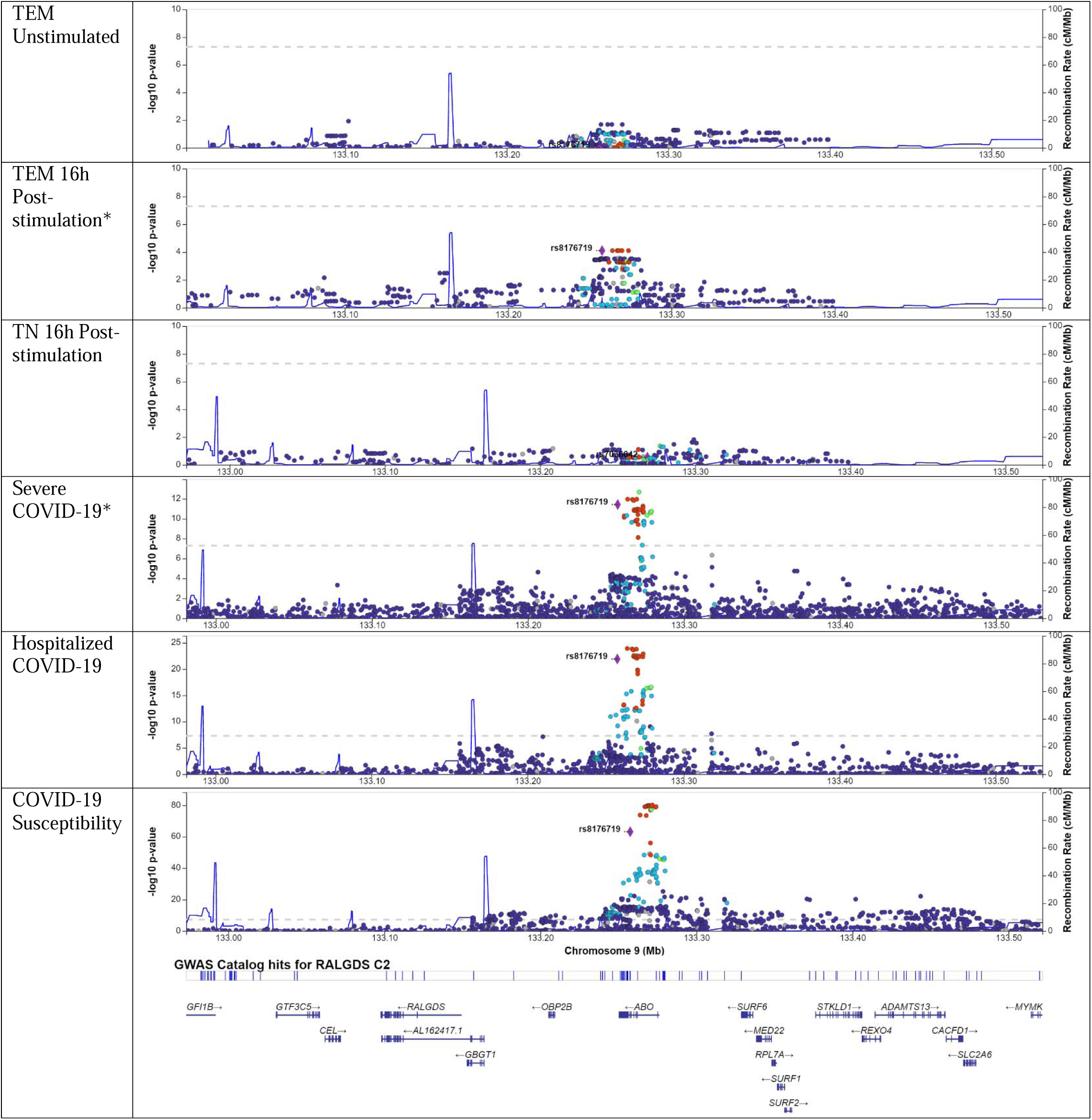
Locuszoom plots for *RALGDS* in the setting of variable cell stimulation and cell type, with selected variant of rs8176719. *RALGDS* was estimated causal with colocalization for only T effectory memory (TEM) cells 16 hours post-stimulation, highlighting the role of cell stimulation on colocalization. A * indicates a dataset that colocalized. rs8176719 was not detected in T naïve cells 16 hours post-stimulation, so rs7036642 was highlighted, which is in LD with rs8176719.

Colocalization of transcripts occurred at varying times following stimulation (**Fig. 2**). Colocalization did not appear to depend on SARS-CoV-2 PCR result in BQC19 among individuals with symptoms of COVID-19, as seen with *IFNAR2* (**Fig. 2, Fig. 5**). Colocalization of select transcripts appeared organ specific, as with *MUC5B* (**Fig. 6**), although there was more noise in bulk sequencing data, as was observed for *ABO.* Specifically, *ABO* colocalized in some organs for only a single outcome and *IL10RB* tested in GTEx had an opposite direction of effect in cultured fibroblasts than tibial nerve (**Fig. 2**).

**Fig 5.**
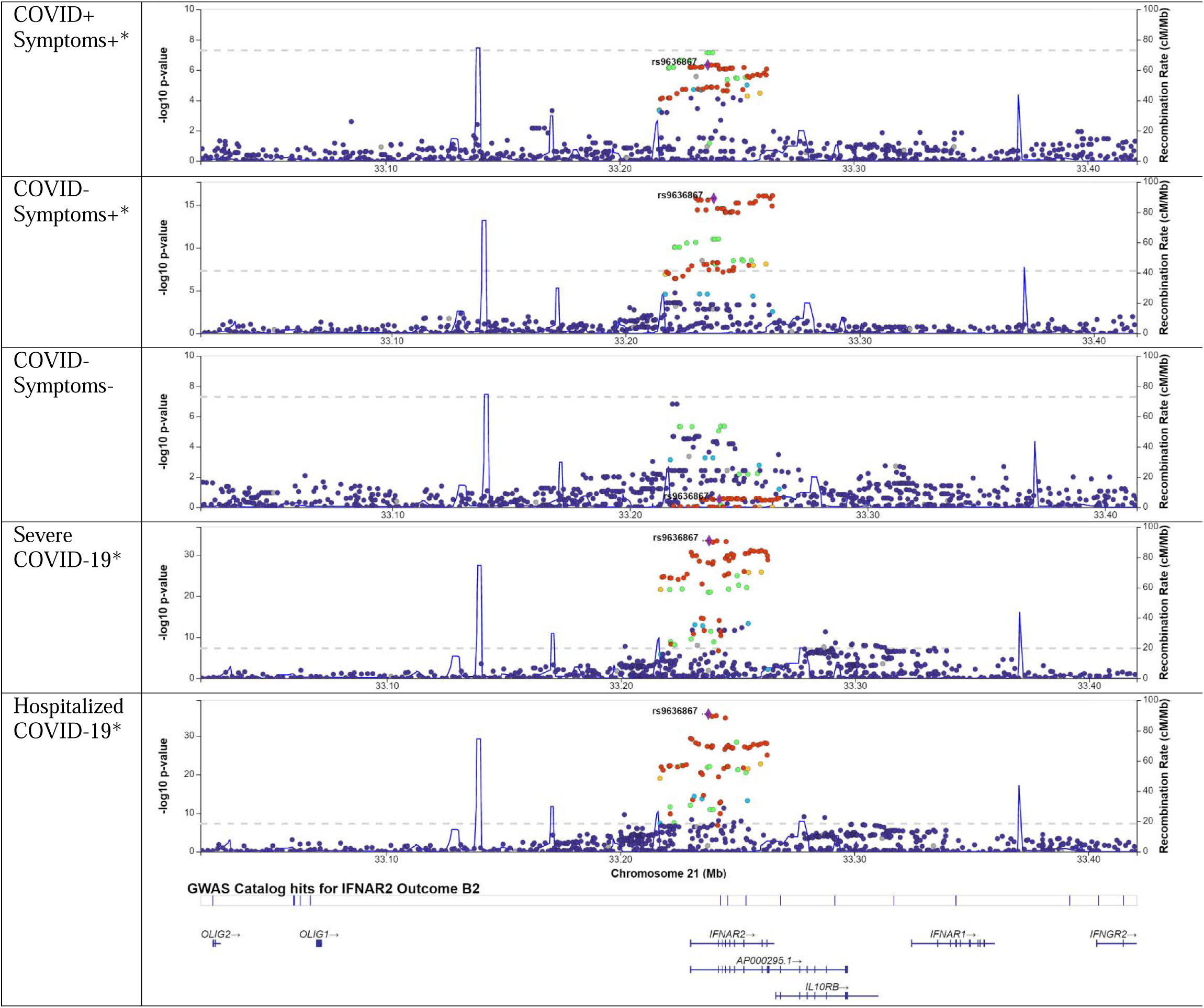
Locuszoom plots for *IFNAR2* in the setting of variable presence of COVID-19 symptoms and SARS-CoV-2 positivity, with selected variant of rs9636867. *IFNAR2* was estimated causal and colocalized only in patients with COVID-19 symptoms (Symptoms +/-) with SARS-CoV-2 status (COVID +/-) having a less apparent role. A * indicates a dataset that colocalized.

**Fig 6.**
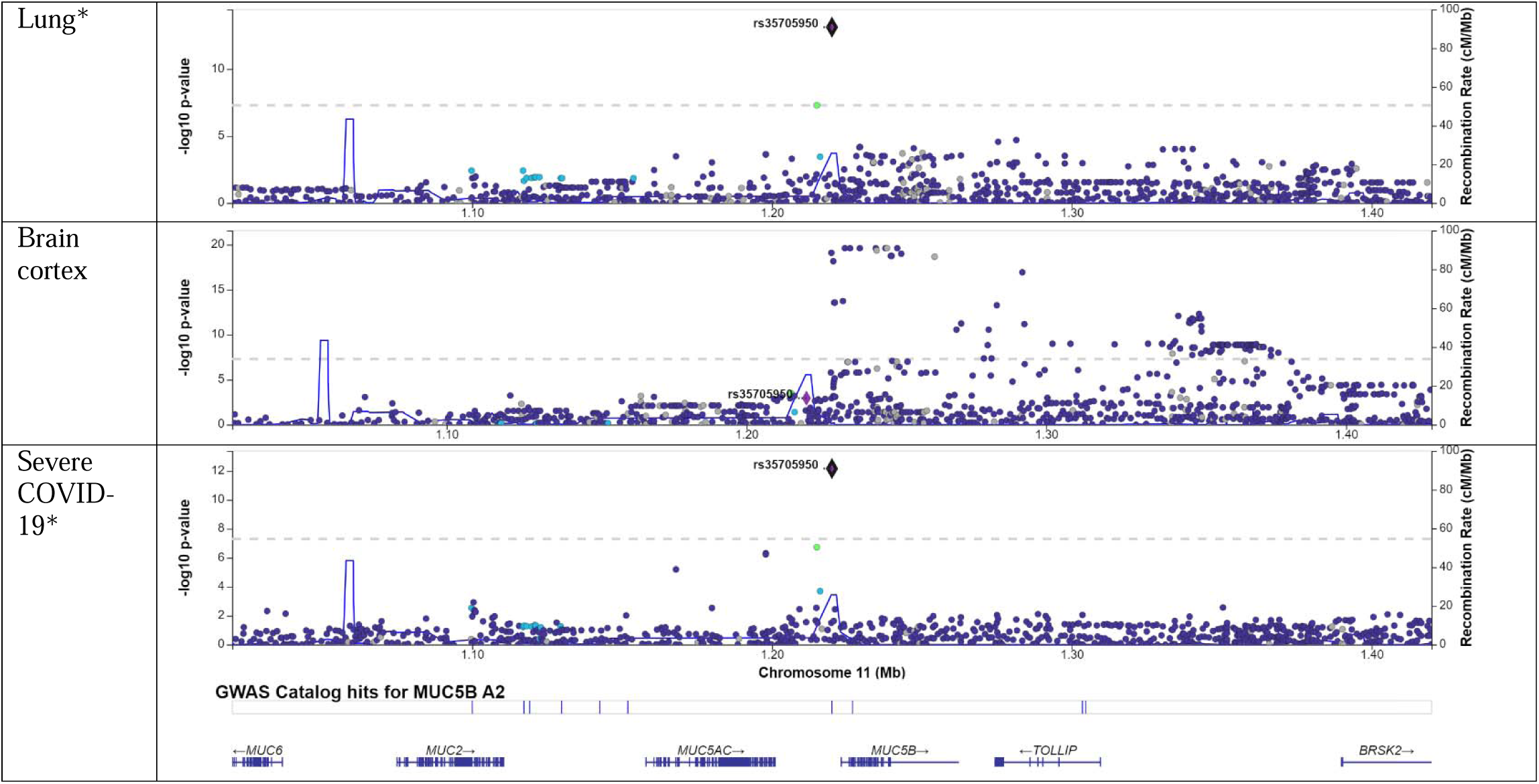
Locuszoom plots for *MUC5B* in the setting of variable organ in GTEx v8, with selected variant of rs35705950. *MUC5B* was estimated causal and colocalized only in lung for severe COVID-19. There were no *cis*- eQTLs for whole blood.

While many transcripts did not colocalize, many transcripts that did have evidence of the single causal variant assumption being violated, with multiple peaks present within or close to the cognate 1 Mb window, suggesting bias due to linkage disequilibrium (**Fig. 7**). Of 19 colocalizing transcripts in single-cell CD4+ T cells, 6 had evidence of violating this assumption. Of 9 colocalizing transcripts in bulk whole blood from patients with symptoms of COVID-19 in BQC19, we observed 6 that violated this assumption. Of 4 from bulk whole blood in GTEx, 2 violators. Of 378 from all tissues in GTEx, 265 transcripts violated this assumption. These results underpin the limitations of present study sample sizes and existing methods designed to clarify causal colocalizing expression signals.

**Fig 7.**
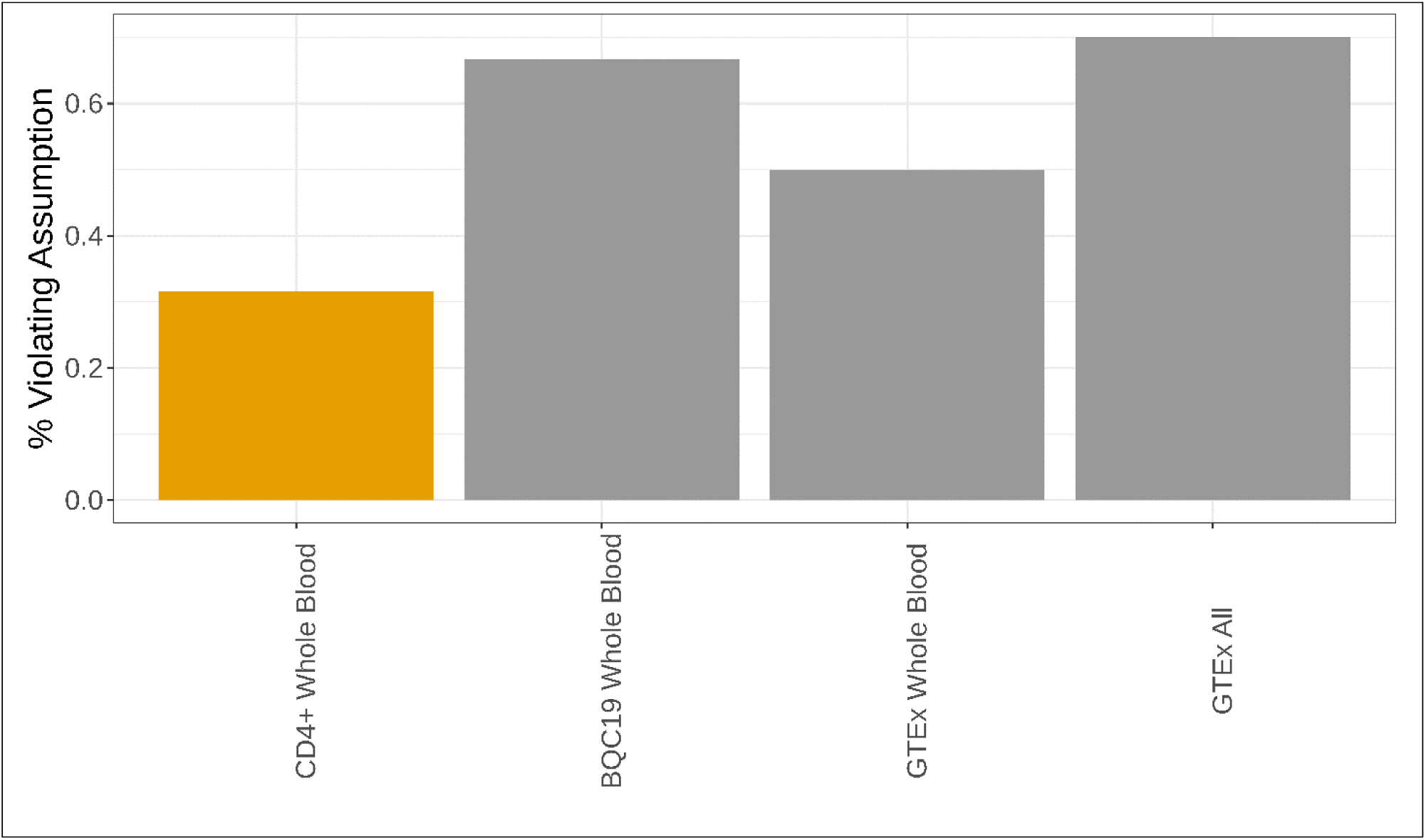
Percentage of colocalizing results (PP_H4_ ≥ 0.80) that had evidence of violating the single causal variant assumption. Orange bars refer to single cell data, gray bulk sequencing.

## Discussion

In this study, we attempted to identify factors that influence colocalization of *cis*-eQTL MR findings for COVID-19 outcomes. Most MR-identified single-cell eQTLs and bulk eQTLs did not colocalize, suggesting that linkage disequilibrium and limited study sizes yielding fewer converging signals may strongly impact the validity of eQTL MR studies that assess colocalization, and necessitate more stringent quality control. Previous research has demonstrated a lack of colocalization of expression findings and suggested that this may be resolved by single-cell eQTL analyses, assessing stimulation state, or clearly defining the state of the individual when blood samples were drawn. Here we show that colocalization of these signals is highly variable, and it is not fully explained by changing cell type, cell stimulation, symptoms of COVID-19, or organ. Taken together, these findings suggest that colocalization of eQTLs with disease outcomes is difficult using current technologies, reference panels and statistical methods, and with present study sample sizes that in single-cell studies typically is limited to 100 individuals.

While we overall found colocalization to be limited, our results were reassuring in that we observed some trends and results consistent with past studies and hypotheses (**Fig. 2**) (Connally et al. 2022; Soskic et al. 2022). For putatively causal transcripts from CD4+ T cells in whole blood, colocalization was influenced by cell stimulation, or cell state (**Fig. 2a**), like other groups’ observations when conducting colocalization without MR (Soskic et al. 2022). On comparing putatively causal transcripts between single-cell and bulk results, with the latter evaluating transcripts in several different cells in a matching tissue, we found no overlap between the modalities in the same tissue, that could suggest that sequencing modality plays a role (**Fig. 2**). Genes that colocalized in CD4+ T cells did not colocalize in spleen that is enriched with T cells (**Fig. 2**). This could underscore the impact of sequencing multiple cell types in bulk sequencing, suggesting that results must be contextualized against sequencing modality and sample cellular heterogeneity. We found organ to play a moderate role in colocalization when comparing putatively causal genes in GTEx (**Fig. 2b**), supporting others’ findings with non-COVID-19 outcomes (Rocheleau et al. 2022). The data on organs’ roles were perhaps noisy. While we found outcome to have a limited role in influencing colocalization for select transcripts, different from others (Rocheleau et al. 2022; Soskic et al. 2022), this could be due to the greater similarity in our outcomes.

This paper has limitations. The scRNA-seq data came from cells stimulated by a standard T cell stimulator rather than SARS-CoV-2. Such stimulation has been employed by several existing works studying immunity in COVID-19 (De Biasi et al. 2020; Kundu et al. 2022; Mathew et al. 2020) and may have some relevance to the stimulation endured when T-cells encounter SARS-CoV-2. We only investigated CD4+ T cells in whole blood (**Fig. 2**). There are other cell types present in greater proportions, which could explain the minimal overlap in colocalization between our single-cell and bulk sequencing results. We could only locate whole blood *cis*-sceQTLs from an admixed study that did not release ancestry-stratified results (Randolph et al. 2021). Ancestry-stratified results are important for Mendelian randomization and colocalization to limit bias from indirect pleiotropic mechanisms, including linkage disequilibrium that varies by ancestral group. Our GTEx results show that many tissues could contribute to COVID-19 outcomes and pathogenesis (**Fig. 2**). Single-cell analysis of all tissues, particularly lung, could help understand the multi-system basis of post-COVID-19 syndrome (Mehandru and Merad 2022). We were unable to access single-cell lung data for this work (Lamontagne et al. 2018). Our outcomes were limited to COVID-19 severity and susceptibility, when expression could also influence complications of COVID-19, such as post-COVID-19 syndrome and schizophrenia (Baranova et al. 2022b; Mehandru and Merad 2022). We have developed two-step MR methods that link gene expression to COVID-19 outcomes, and then to these complications (Yoshiji et al. 2023), which has already been used to validate BMI’s role on COVID-19 outcomes as discovered by others (Baranova et al. 2023).

Our study was limited to individuals of European ancestry. While the HGI has found loci with genome-significant variants in other ancestries, none of the loci from Admixed American, African, East Asian, or South Asian ancestry for any outcome overlapped with loci found to influence COVID-19 outcomes in this study. Given fewer loci in non-European datasets, this could be due to sample size and underscores the importance of multi-ancestry analyses. However, even within European ancestry individuals, subtle differences in LD patterns can influence colocalization, which we observed (**Fig. 4-6**) (Kanai et al. 2022; Kanai et al. 2021). Such differences may have impacted the lack of colocalization observed here. Sample size was limited for the eQTL datasets that we employed, which increases the risk of biased results, which we possibly observed where some genes colocalized for only a single outcome or did not colocalize in one outcome (**Fig. 2**). We adjusted for this by using methods well established in the literature, including several stringent and conservative sensitivity tests and means of quality control for MR and colocalization. While using a colocalization window around a lead variant of ±500 kb is more likely to limit bias from pleiotropy, it increases the risk of missed signals. Data must still be carefully appraised for possible false positives, as may be the case with *IL10RB* being estimated protective in cultured fibroblasts for all outcomes but increasing the risk of COVID-19 outcomes in the tibial nerve for all outcomes (**Fig. 2**).

While we found symptoms of COVID-19 to influence genomic colocalization for select transcripts and states, as shown by *IFNAR2* between severe and hospitalized COVID-19, this trend could have been influenced by a batch effect. SARS-CoV-2 status’ effect on colocalization among individuals in BQC19 could be biased as negative results could represent false negatives and individuals with COVID-19 symptoms could have had a different viral illness. We mitigated this by deriving eQTLs using the same pipeline used by GTEx and limiting the conclusions we made given this context (GTEx Consortium 2020). Generally, we investigated variables in multiple datasets, given that a variable’s demonstrated role in multiple cohorts supports making a generalization.

## Conclusions

While existing hypotheses suggest that colocalization of transcripts depends on multiple conditions, we found that *cis*-eQTLs identified by MR for COVID-19 outcomes rarely colocalized, even when assessing different cell types, cell states, symptoms of COVID-19 and organs. Taken together, these findings suggest that even after accounting for variables, there was little evidence of colocalization for most genes whose influence on COVID-19 outcomes was identified through MR.

## Data Availability

Data from Soskic et al. are available through Zenodo (doi:10.5281/zenodo.6006796). BQC-19 data, including expression data, is available by application: https://www.bqc19.ca/. GTEx release 8 data is available from: https://www.gtexportal.org/home/. COVID-19 HGI summary statistics are available from: https://www.covid19hg.org/.

## Supplemental Table Captions

**Supplemental Table 1.** All lead variants in HGI European-ancestry data, excluding MHC loci.

**Supplemental Table 2.** All genes that were estimated causal and colocalized in our study, with existing evidence linking genes to outcomes.

| Gene | Colocalizing cell/tissue | Existing evidence |
| --- | --- | --- |
| *ABO* | Adrenal gland (Severe) - Hazardous<br>Artery tibial (Severe) – Hazardous<br>Breast Mammary Tissue (Susceptible) – Hazardous<br>Esophagus mucosa (Susceptible) - Hazardous<br>Esophagus muscularis (Severe) - Hazardous<br>Heart Left Ventricle (Severe and Hospitalized) – Hazardous<br>Liver (Hospitalized) - Hazardous<br>Muscle skeletal (Severe) - Hazardous<br>Testis (All) - Hazardous<br>Whole blood COVID+ Symptoms+ (Hospitalized) - **Protective** | MR in non-GTEx and GTEx eQTL and pQTL datasets for lung and whole blood (Baranova et al. 2022a; Hernandez Cordero et al. 2021) |
| *ADAM15* | T central memory Stim 5d (Susceptible) - Hazardous | MR with colocalization in GTEx did not observe colocalization (Wang et al. 2022) |
| *ATP11A* | Artery Aorta (Hospitalized) - Protective | Fine mapping (Kousathanas et al. 2022) |
| *ATP5PO* | Lung (Hospitalized) - Hazardous | Associational (Li et al. 2022) |
| *CNPY4* | Testis (Severe) - Protective | None |
| *COPS6* | Esophagus muscularis (Hospitalized) - Protective | None |
| *DLX3* | Lung (Hospitalized) - Hazardous | MR with colocalization in lung (Hernandez Cordero et al. 2021) |
| *DPP9* | Skin sun exposed lower leg (All) - Protective | Associational data in blood (Sharif-Zak et al. 2022) |
| *FAM55C* | Esophagus muscularis (Susceptible) - Hazardous | None |
| *GBA* | Nerve Tibial (Susceptible) - Hazardous<br>Thyroid (Susceptible) - Hazardous | None |
| *GBAP1* | Breast Mammary Tissue (Susceptible) - Hazardous<br>Cultured Fibroblasts (Susceptible) - Hazardous<br>Colon Sigmoid (Susceptible) - Hazardous<br>Lung (Susceptible) - Hazardous<br>Nerve Tibial (Susceptible) - Hazardous<br>Small Intestine Terminal Ileum (Susceptible) - Hazardous<br>Stomach (Susceptible) - Hazardous | MR with colocalization for splice isoform (Nakanishi et al. 2022) |
| *HIP1* | Breast mammary tissue (Severe) - Hazardous<br>Nerve tibial (Severe) - Hazardous<br>Whole blood COVID- Symptoms- (Severe) - **Protective** | None |
| *IFNAR2* | Cells cultured fibroblasts (Severe and hospitalized) - Protective<br>Esophagus mucosa (Susceptible) - Protective<br>Whole blood COVID- Symptoms+ (Hospitalized) - Protective<br>Whole blood COVID+ Symptoms+ (Severe) - Protective | MR with colocalization for GTEx v.7 eQTLs and pQTLs for whole-blood and lung (Baranova et al. 2021; Fricke-Galindo et al. 2022; Krishnamoorthy et al. 2023; Liu et al. 2021; Pairo-Castineira et al. 2021) |
| *IL10RB* | Cells Cultured fibroblasts (All) - **Protective**<br>Esophagus Gastroesophageal junction (All) - Hazardous<br>Skeletal muscle (Susceptible) - Hazardous<br>Nerve tibial (All) - Hazardous | MR with colocalization for eQTLs and pQTLs, did not study excitatory neurons (Gaziano et al. 2021) |
| *KCNC3* | Tibial nerve (Severe and Hospitalized) – Hazardous | TWAS (Krishnamoorthy et al. 2023) |
| *LINC01301* | Breast mammary tissue (Hospitalized) – Protective<br>Skin not sun exposed (Hospitalized) - Protective | TWAS (Pairo-Castineira et al. 2022) |
| *LINC02210* | Artery tibial (Severe) – Protective | MR with colocalization (Baranova et al. 2021) |
| *Lnc-INTS12-1* | Artery aorta (Severe) – Protective<br>Thyroid (Severe) - Protective | None |
| *Lnc-THBS3-1* | Whole blood COVID- Symptoms- (Susceptible) - Protective | None |
| *MAMSTR* | Skin Sun Exposed (Hospitalized) – Protective | TWAS (Krishnamoorthy et al. 2023) |
| *MUC1* | Nerve tibial (Severe and Hospitalized) - Hazardous | MR from GTEx v.7 against HGI release 6 found signal in blood (Krishnamoorthy et al. 2023) |
| *MUC5B* | Lung (Severe) - Protective | MR only for lung eQTLs GTEx release 7 (Liu et al. 2021) |
| *NAPSA* | CD4 Naïve Stim 40h (Susceptible) - Hazardous<br>Esophageal mucosa (Severe and Hospitalized) - **<u>Protective</u>**<br>T effector memory Stim 40h (All) - Hazardous<br>T naïve (Hospitalized and Susceptible) - Hazardous<br>Testis (Hospitalized) - **<u>Protective</u>** | MR with colocalization for eQTLs, did not study CD4+ T cells (Degenhardt et al. 2022) |
| *NAPSB* | Artery Coronary (Susceptible) - Protective | None |
| *NTN5* | Colon sigmoid (Hospitalized) - Protective<br>Esophageal Gastroesophageal junction (Hospitalized) - Protective<br>Esophagus mucosa (Severe and Hospitalized) - Protective | None |
| *OAS1* | Adipose subcutaneous (Hospitalized) - Protective<br>Artery tibial (Susceptible) - Protective<br>Esophagus mucosa (Hospitalized) - **<u>Hazardous</u>**<br>Nerve tibial (Susceptible) - **<u>Hazardous</u>**<br>Skin not sun exposed (Severe and Susceptible) - Protective<br>Skin sun exposed (All) - Protective | MR with colocalization of eQTL and pQTL (Zhou et al. 2021) |
| *PDCL3P4* | Brain Caudate Basal Ganglia (Susceptible) | Colocalization (D'Antonio et al. 2021) |
| *RAB2A* | Adipose subcutaneous (Hospitalized) - Hazardous<br>Artery aorta (Hospitalized) - Hazardous<br>Artery tibial (Hospitalized) - Hazardous<br>CD4 Memory Stim 40h (Hospitalized) - Hazardous<br>CD4 Naïve Stim 16h (Hospitalized) - Hazardous<br>CD4 Naïve Stim 40h (Hospitalized) - Hazardous<br>Esophagus mucosa (Hospitalized) - Hazardous<br>Esophagus muscularis (Hospitalized) - Hazardous<br>Liver (Hospitalized) - Hazardous<br>Lung (Hospitalized) - Hazardous<br>Skin not sun exposed (Hospitalized) - Hazardous<br>T naïve Stim 40h (Hospitalized) - Hazardous | MR and colocalization for pQTLs, but not for eQTLs (Pietzner et al. 2022) |
| *RALGDS* | T effect memory Stim 16h (Severe and Susceptible) - Protective | None |
| *RASIP1* | Esophageal Gastroesophageal junction (Severe and Hospitalized) - Protective<br>Esophagus muscularis (Severe) - Protective | None |
| *SLC22A31* | Heart left ventricle (Severe and Hospitalized) – Hazardous | PheWAS linked (Covid-19 Host Genetics Initiative 2022) |
| *SLC6A20* | Breast mammary tissue (Severe and Hospitalized) – Hazardous | GWAS (Severe Covid et al. 2020) |
| *WNT3* | Adrenal Gland (Hospitalized) - Hazardous<br>Artery Aorta (Severe and Hospitalized) - **<u>Protective</u>**<br>Colon Transverse (Severe) - Hazardous<br>Nerve Tibial (Severe) - Hazardous<br>Thyroid (Severe and Hospitalized) - Hazardous | Fine mapped (Wu et al. 2021) |

**Supplemental Table 3**. All MR results using CD4+ T cell expression data.

**Supplemental Table 4**. All MR results using expression data from individuals with symptoms of COVID-19 in BQC19.

**Supplemental Table 5**. All MR results using expression data from all tissues in GTEx.

**Supplemental Table 6**. All colocalization results of putatively causal variants from CD4+ T cell expression data.

**Supplemental Table 7**. All colocalization results of putatively causal variants from individuals with symptoms of COVID-19 in BQC19.

**Supplemental Table 8**. All colocalization results of putatively causal variants from all tissues in GTEx.

## Methods

### Datasets

We examined *cis*-eQTLs in three datasets to implicate their influence on COVID-19 outcomes and investigate how experimental and physiological conditions impact colocalization (**Fig. 1**). scRNA-seq *cis*-eQTLs from Soskic *et al*. were used to analyze how cell type, cell stimulation, and cell microenvironment affected colocalization (Soskic et al. 2022). Bulk RNA-seq *cis*-eQTLs from Biobanque Quebecoise de la COVID-19 (BQC19) were used to investigate how disease state impacted identified *cis*-eQTLs and colocalization in individuals with symptoms of COVID-19, with and without PCR-confirmed SARS-CoV-2 infection. Bulk RNA-seq *cis*-eQTLs from GTEx whole blood were used to compare BQC19 data with data from individuals without symptoms of COVID-19 with all other organs used to determine the role of organ (GTEx Consortium 2020).

#### Whole-blood single-cell eQTLs

The summary statistics for immune cell expression *cis*-eQTLs before and after stimulation with anti-CD3/anti-CD28 human T-Activators were obtained from Soskic *et al*. (Soskic et al. 2022). The study consisted of 119 individuals of British ancestry with peripheral blood mononuclear cells (655,349 CD4+ T cells) sequenced using scRNA-seq (Soskic et al. 2022). We used the summary statistics for cells at all available time points (unstimulated, 16 hours post-stimulation corresponding to before cell division, 40 hours post-stimulation corresponding to after cell division, five days post-stimulation corresponding to gaining effector function) for cell types present before stimulation and present for at least one time point after stimulation (Soskic et al. 2022). The unstimulated time point acted as a control for stimulation, sequenced 16 hours after culturing without any anti- CD3/anti-CD28 human T-Activators (Soskic et al. 2022). We investigated CD4+ antigen-I and CD4+ memory cell classifications before Leiden-algorithm clustering, implemented by Soskic et al., and T I, T central memory, T effector memory, CD45RA re-expressing T effector memory, and thymus-derived regulatory T cells after clustering (Soskic et al. 2022). Full details describing RNA sequencing, separation of cell types and stimulation are available in Soskic *et al*. (Soskic et al. 2022).

#### Bulk whole blood eQTLs from subjects with symptoms of COVID-19, with and without SARS-CoV-2 positive PCR results

BQC19 (https://en.quebeccovidbiobank.ca) is a prospective cohort enrolling participants with PCR-proven SARS-CoV-2 infection and PCR-proven SARS-CoV-2 negative individuals who presented to the hospital with signs or symptoms consistent with COVID-19. Participants were recruited from eight academic hospitals in the province of Quebec, Canada. A total of 4,704 participants underwent PCR testing with confirmed positive or negative results between January 25^th^, 2020 to March 20^th^, 2022. A total of 379 participants had RNA extracted and sequenced with eQTLs called using the GTEx pipeline, available from the Broad Institute (https://github.com/broadinstitute/gtex-pipeline/tree/master/qtl). We used data solely for those of non-Finnish European ancestry, which included 112 SARS-CoV-2 positive and 166 SARS-CoV-2 negative samples.

#### GTEx release 8 data

We obtained *cis*-eQTL summary statistics for GTEx release 8 for whole-blood and all other organs, restricted to individuals of European ancestry, from: https://www.gtexportal.org/home/ (GTEx Consortium 2020).

#### COVID-19 Outcome Data

European-ancestry-specific summary statistics for COVID-19 outcomes were obtained from the COVID-19 Host Genetics Initiative (HGI) release 7 (https://www.covid19hg.org/), which did not include individuals from 23AndMe (Covid-19 Host Genetics Initiative 2021). The COVID-19 outcomes included severe COVID-19 (13,769 cases and 1,072,442 controls), COVID-19 hospitalization (32,519 cases and 2,062,805 controls), and susceptibility to COVID-19 (122,616 cases and 2,475,240 controls). Severe COVID-19 was defined as COVID- 19 requiring respiratory support or resulting in death. COVID-19 with hospitalization was defined as an infection requiring hospitalization or death. COVID-19 susceptibility was defined as infection determined by self-report on a questionnaire, electronic medical record diagnosis, or laboratory testing (serology tests or nucleic acid amplification). Controls were all individuals who did not meet an outcome’s definition.

### MR of *cis*-eQTLs with COVID-19 outcomes

We used MR to estimate the causal relationship between exposures (which here are RNA transcript levels) and COVID-19 outcomes to determine how cell type, cell stimulation, time after stimulation, symptoms of COVID-19, PCR result for SARS-CoV-2 among individuals with symptoms of COVID-19, and organ influenced the relationship between gene expression and COVID-19 outcomes. MR studies use SNPs strongly associated with an exposure as instrumental variables to estimate the effect of an exposure on an outcome. Such studies reduce potential confounding effects because genetic variants are essentially randomized at conception. They also prevent bias due to reverse causation (wherein the outcome influences the exposure) since the assignment of genetic variants always precedes disease onset (Smith and Ebrahim 2003). The main assumptions of MR are that the variants under study are strongly associated with the risk factor of interest, confounders of the exposure-outcome relationship are not associated with the variants, and the variants only affect the outcome through the risk factor (Davies et al. 2018; Skrivankova et al. 2021). The most problematic of these assumptions is the last, as it is difficult to confidently understand if the SNPs affect the outcome independent of the exposure (i.e. a lack of horizontal pleiotropy). To partially mitigate against such potential bias, we have used only *cis*-eQTLs, which are more likely to act directly through the transcription or translation of the proximal gene, rather than through horizontally pleiotropic pathways. There was overlap of individuals from the *cis*-eQTL studies with individuals in the HGI data.

To undertake MR analyses, we used the TwoSampleMR (v0.5.6) package (https://mrcieu.github.io/TwoSampleMR/). First, we identified all genome significant (p ≤ 5 x 10^-8^) loci from HGI COVID-19 summary statistics. We isolated all variants 500 kb upstream and downstream of the lead HGI variant for each locus for each exposure and outcome. Second, we excluded the MHC locus (chr6: 28,510,120 – 33,480,577; GRCh38) to reduce potential confounding by linkage disequilibrium structure. Third, the exposure *cis*-eQTLs, ±500 kb from their transcriptional start sites, for each cell type, cell stimulation state, cell stimulation time, SARS-CoV-2 status, presence of COVID-19 symptoms, and gene were filtered for variants in LD using the package’s clumping function with default settings. Fourth, the exposure data were harmonized with outcome data using default settings. Fifth, we conducted MR with sensitivity tests. For loci with only one remaining *cis*-eQTL following harmonization, we applied Wald Ratio-based MR. For loci with more than one remaining independent *cis* variant, we used inverse-variance weighted MR (MR-IVW). For loci with three or more remaining *cis*-eQTLs, we did sensitivity testing using exposure MR weighted median, MR weighted penalized median, MR weighted mode, MR Egger regression, and MR Egger intercept. MR Steiger was done for every test to assess for reverse causation, wherein the MR results would be better explained by the effects of COVID-19 on the *cis*-eQTL. Results were retained for downstream analysis if they passed these sensitivity tests with a Wald Ratio or IVW Benjamini-Hochberg adjusted p-value, which controls the false-discovery rate by adjusting the p-value by the number of tests, less than or equal to 0.05.

### Colocalization

To investigate whether *cis*-eQTLs shared the same single causal signal with COVID-19 outcomes, we used coloc v5.1.1 (https://chr1swallace.github.io/coloc/). Colocalization helps to guard against bias due to confounding from linkage disequilibrium. Such confounding can occur when the SNPs that influence an exposure (here *cis*-eQTLs) do not causally influence an outcome (here COVID-19 outcomes) but are associated with each other due to linkage disequilibrium. Consistent with Soskic *et al*., we required at least 50 variants for each colocalization analysis (Soskic et al. 2022). We employed default priors of p_1_=p_2_=10^-4^ and p_12_=10^-5^ where p_1_ is the prior probability that only eQTLs had a genetic association in the region, p_2_ is a prior probability that only the HGI summary statistics had a genetic association in the region, and p_12_ is the prior probability that the eQTL data and the HGI summary statistics shared the same genetic associations in the region. A p_12_ > 0.8, or PP_H4_ > 0.8 was considered evidence of colocalization. Sensitivity tests were conducted using coloc’s sensitivity test function for each instance of colocalization to validate the single causal variant assumption and evaluate the robustness of results to different prior settings. Locuszoom plots used to highlight trends were produced using Locuszoom (Pruim et al. 2010).

### Ethics statement

Soskic *et al*. biological samples were ethically sourced and used in research under an institutional review board-approved protocol (Soskic et al. 2022). BQC19 received ethical approval from the Jewish General Hospital research ethics board (2020-2137) and the Centre Hospitalier de l’Université de Montréal institutional ethics board (MP-02-2020-8929, 19.389). Specimens collected by GTEx were sourced per protocol for an institutional review board (GTEx Consortium 2020).

### Data availability

Data from Soskic *et al*. are available through Zenodo (doi:10.5281/zenodo.6006796). BQC-19 data, including expression data, is available by application: https://www.bqc19.ca/. GTEx release 8 data is available from: https://www.gtexportal.org/home/. COVID-19 HGI summary statistics are available from: https://www.covid19hg.org/.

### Code availability

All analyses were conducted in R v4.0.5 (R Core Team 2021). Code is available at: https://github.com/richardslab/dynamic_QTL_COVID19.

## Acknowledgements

JDSW is a graduate student in McGill’s Quantitative Life Sciences Ph.D. program. This work was made possible through open sharing of data and samples from BQC19. We thank all participants of BQC19 for their contribution. We thank Vincent Mooser, director of BQC19, for his contributions organizing BQC19. Some transcriptomics sequencing was performed at the McGill University Genome Centre (Montreal, Quebec, Canada) by Ariane Boisclair, Janick St-Cyr, Pouria Jandaghi, Daniel Auld. The remainder of the transcriptomics sequencing was performed at the Centre d’expertise et de services Génome Québec (Montreal, Quebec, Canada), facilitated by the diligent work of Alexandre Montpetit, Isabelle Guillet, Joelle Fontaine, Martine Lavoie, and Genevieve DonPierre. Kevin Liang contributed to paper revisions.

## References

Baranova A, Cao H, Chen J, Zhang F (2022a) Causal Association and Shared Genetics Between Asthma and COVID-19. Front Immunol 13: 705379. doi: 10.3389/fimmu.2022.705379

Baranova A, Cao H, Teng S, Zhang F (2023) A phenome-wide investigation of risk factors for severe COVID-19. J Med Virol 95: e28264. doi: 10.1002/jmv.28264

Baranova A, Cao H, Zhang F (2021) Unraveling Risk Genes of COVID-19 by Multi-Omics Integrative Analyses. Front Med (Lausanne) 8: 738687. doi: 10.3389/fmed.2021.738687

Baranova A, Cao H, Zhang F (2022b) Severe COVID-19 increases the risk of schizophrenia. Psychiatry Res 317: 114809. doi: 10.1016/j.psychres.2022.114809

Connally NJ, Nazeen S, Lee D, Shi H, Stamatoyannopoulos J, Chun S, Cotsapas C, Cassa CA, Sunyaev SR (2022) The missing link between genetic association and regulatory function. Elife 11. doi: 10.7554/eLife.74970

Covid-19 Host Genetics Initiative (2021) Mapping the human genetic architecture of COVID-19. Nature 600: 472–477. doi: 10.1038/s41586-021-03767-x

Covid-19 Host Genetics Initiative (2022) A first update on mapping the human genetic architecture of COVID-19. Nature 608: E1–E10. doi: 10.1038/s41586-022-04826-7

D’Antonio M, Nguyen JP, Arthur TD, Matsui H, Initiative C-HG, D’Antonio-Chronowska A, Frazer KA (2021) SARS-CoV-2 susceptibility and COVID-19 disease severity are associated with genetic variants affecting gene expression in a variety of tissues. Cell Rep 37: 110020. doi: 10.1016/j.celrep.2021.110020

Davies NM, Holmes MV, Davey Smith G (2018) Reading Mendelian randomisation studies: a guide, glossary, and checklist for clinicians. BMJ 362: k601. doi: 10.1136/bmj.k601

De Biasi S, Meschiari M, Gibellini L, Bellinazzi C, Borella R, Fidanza L, Gozzi L, Iannone A, Lo Tartaro D, Mattioli M, Paolini A, Menozzi M, Milic J, Franceschi G, Fantini R, Tonelli R, Sita M, Sarti M, Trenti T, Brugioni L, Cicchetti L, Facchinetti F, Pietrangelo A, Clini E, Girardis M, Guaraldi G, Mussini C, Cossarizza A (2020) Marked T cell activation, senescence, exhaustion and skewing towards TH17 in patients with COVID-19 pneumonia. Nat Commun 11: 3434. doi: 10.1038/s41467-020-17292-4

Degenhardt F, Ellinghaus D, Juzenas S, Lerga-Jaso J, Wendorff M, Maya-Miles D, Uellendahl-Werth F, ElAbd H, Ruhlemann MC, Arora J, Ozer O, Lenning OB, Myhre R, Vadla MS, Wacker EM, Wienbrandt L, Blandino Ortiz A, de Salazar A, Garrido Chercoles A, Palom A, Ruiz A, Garcia-Fernandez AE, Blanco-Grau A, Mantovani A, Zanella A, Holten AR, Mayer A, Bandera A, Cherubini A, Protti A, Aghemo A, Gerussi A, Ramirez A, Braun A, Nebel A, Barreira A, Lleo A, Teles A, Kildal AB, Biondi A, Caballero-Garralda A, Ganna A, Gori A, Gluck A, Lind A, Tanck A, Hinney A, Carreras Nolla A, Fracanzani AL, Peschuck A, Cavallero A, Dyrhol-Riise AM, Ruello A, Julia A, Muscatello A, Pesenti A, Voza A, Rando-Segura A, Solier A, Schmidt A, Cortes B, Mateos B, Nafria-Jimenez B, Schaefer B, Jensen B, Bellinghausen C, Maj C, Ferrando C, de la Horra C, Quereda C, Skurk C, Thibeault C, Scollo C, Herr C, Spinner CD, Gassner C, Lange C, Hu C, Paccapelo C, Lehmann C, Angelini C, Cappadona C, Azuure C, Covicat study group AS, Bianco C, Cea C, Sancho C, Hoff DAL, Galimberti D, Prati D, Haschka D, Jimenez D, Pestana D, Toapanta D, Muniz-Diaz E, Azzolini E, Sandoval E, Binatti E, Scarpini E, Helbig ET, et al. (2022) Detailed stratified GWAS analysis for severe COVID-19 in four European populations. Hum Mol Genet 31: 3945–3966. doi: 10.1093/hmg/ddac158

Forgetta V, Jiang L, Vulpescu NA, Hogan MS, Chen S, Morris JA, Grinek S, Benner C, Jang DK, Hoang Q, Burtt N, Flannick JA, McCarthy MI, Fauman E, Greenwood CMT, Maurano MT, Richards JB (2022) An effector index to predict target genes at GWAS loci. Hum Genet. doi: 10.1007/s00439-022-02434-z

Fricke-Galindo I, Martinez-Morales A, Chavez-Galan L, Ocana-Guzman R, Buendia-Roldan I, Perez- Rubio G, Hernandez-Zenteno RJ, Veronica-Aguilar A, Alarcon-Dionet A, Aguilar-Duran H, Gutierrez-Perez IA, Zaragoza-Garcia O, Alanis-Ponce J, Camarena A, Bautista-Becerril B, Nava- Quiroz KJ, Mejia M, Guzman-Guzman IP, Falfan-Valencia R (2022) IFNAR2 relevance in the clinical outcome of individuals with severe COVID-19. Front Immunol 13: 949413. doi: 10.3389/fimmu.2022.949413

Gaziano L, Giambartolomei C, Pereira AC, Gaulton A, Posner DC, Swanson SA, Ho YL, Iyengar SK, Kosik NM, Vujkovic M, Gagnon DR, Bento AP, Barrio-Hernandez I, Ronnblom L, Hagberg N, Lundtoft C, Langenberg C, Pietzner M, Valentine D, Gustincich S, Tartaglia GG, Allara E, Surendran P, Burgess S, Zhao JH, Peters JE, Prins BP, Angelantonio ED, Devineni P, Shi Y, Lynch KE, DuVall SL, Garcon H, Thomann LO, Zhou JJ, Gorman BR, Huffman JE, O’Donnell CJ, Tsao PS, Beckham JC, Pyarajan S, Muralidhar S, Huang GD, Ramoni R, Beltrao P, Danesh J, Hung AM, Chang KM, Sun YV, Joseph J, Leach AR, Edwards TL, Cho K, Gaziano JM, Butterworth AS, Casas JP, Initiative VAMVPC-S (2021) Actionable druggable genome-wide Mendelian randomization identifies repurposing opportunities for COVID-19. Nat Med 27: 668–676. doi: 10.1038/s41591-021-01310-z

GTEx Consortium (2020) The GTEx Consortium atlas of genetic regulatory effects across human tissues. Science 369: 1318–1330. doi: 10.1126/science.aaz1776

Hernandez Cordero AI, Li X, Milne S, Yang CX, Bosse Y, Joubert P, Timens W, van den Berge M, Nickle D, Hao K, Sin DD (2021) Multi-omics highlights ABO plasma protein as a causal risk factor for COVID-19. Hum Genet 140: 969–979. doi: 10.1007/s00439-021-02264-5

Kanai M, Elzur R, Zhou W, Global Biobank Meta-analysis I, Daly MJ, Finucane HK (2022) Meta- analysis fine-mapping is often miscalibrated at single-variant resolution. Cell Genom 2. doi: 10.1016/j.xgen.2022.100210

Kanai M, Ulirsch JC, Karjalainen J, Kurki M, Karczewski KJ, Fauman E, Wang QS, Jacobs H, Aguet F, Ardlie KG, Kerimov N, Alasoo K, Benner C, Ishigaki K, Sakaue S, Reilly S, Kamatani Y, Matsuda K, Palotie A, Neale BM, Tewhey R, Sabeti PC, Okada Y, Daly MJ, Finucane HK (2021) Insights from complex trait fine-mapping across diverse populations. medRxiv. doi: 10.1101/2021.09.03.21262975

Kousathanas A, Pairo-Castineira E, Rawlik K, Stuckey A, Odhams CA, Walker S, Russell CD, Malinauskas T, Wu Y, Millar J, Shen X, Elliott KS, Griffiths F, Oosthuyzen W, Morrice K, Keating S, Wang B, Rhodes D, Klaric L, Zechner M, Parkinson N, Siddiq A, Goddard P, Donovan S, Maslove D, Nichol A, Semple MG, Zainy T, Maleady-Crowe F, Todd L, Salehi S, Knight J, Elgar G, Chan G, Arumugam P, Patch C, Rendon A, Bentley D, Kingsley C, Kosmicki JA, Horowitz JE, Baras A, Abecasis GR, Ferreira MAR, Justice A, Mirshahi T, Oetjens M, Rader DJ, Ritchie MD, Verma A, Fowler TA, Shankar-Hari M, Summers C, Hinds C, Horby P, Ling L, McAuley D, Montgomery H, Openshaw PJM, Elliott P, Walsh T, Tenesa A, Gen Oi, and Me i, Initiative C-HG, Fawkes A, Murphy L, Rowan K, Ponting CP, Vitart V, Wilson JF, Yang J, Bretherick AD, Scott RH, Hendry SC, Moutsianas L, Law A, Caulfield MJ, Baillie JK (2022) Whole-genome sequencing reveals host factors underlying critical COVID-19. Nature 607: 97–103. doi: 10.1038/s41586-022-04576-6

Krishnamoorthy S, Li GH, Cheung CL (2023) Transcriptome-wide summary data-based Mendelian randomization analysis reveals 38 novel genes associated with severe COVID-19. J Med Virol 95: e28162. doi: 10.1002/jmv.28162

Kundu R, Narean JS, Wang L, Fenn J, Pillay T, Fernandez ND, Conibear E, Koycheva A, Davies M, Tolosa-Wright M, Hakki S, Varro R, McDermott E, Hammett S, Cutajar J, Thwaites RS, Parker E, Rosadas C, McClure M, Tedder R, Taylor GP, Dunning J, Lalvani A (2022) Cross-reactive memory T cells associate with protection against SARS-CoV-2 infection in COVID-19 contacts. Nat Commun 13: 80. doi: 10.1038/s41467-021-27674-x

Lamontagne M, Berube JC, Obeidat M, Cho MH, Hobbs BD, Sakornsakolpat P, de Jong K, Boezen HM, International CGC, Nickle D, Hao K, Timens W, van den Berge M, Joubert P, Laviolette M, Sin DD, Pare PD, Bosse Y (2018) Leveraging lung tissue transcriptome to uncover candidate causal genes in COPD genetic associations. Hum Mol Genet 27: 1819–1829. doi: 10.1093/hmg/ddy091

Li P, Ke Y, Shen W, Shi S, Wang Y, Lin K, Guo X, Wang C, Zhang Y, Zhao Z (2022) Targeted screening of genetic associations with COVID-19 susceptibility and severity. Front Genet 13: 1073880. doi: 10.3389/fgene.2022.1073880

Liu D, Yang J, Feng B, Lu W, Zhao C, Li L (2021) Mendelian randomization analysis identified genes pleiotropically associated with the risk and prognosis of COVID-19. J Infect 82: 126–132. doi: 10.1016/j.jinf.2020.11.031

Mathew D, Giles JR, Baxter AE, Oldridge DA, Greenplate AR, Wu JE, Alanio C, Kuri-Cervantes L, Pampena MB, D’Andrea K, Manne S, Chen Z, Huang YJ, Reilly JP, Weisman AR, Ittner CAG, Kuthuru O, Dougherty J, Nzingha K, Han N, Kim J, Pattekar A, Goodwin EC, Anderson EM, Weirick ME, Gouma S, Arevalo CP, Bolton MJ, Chen F, Lacey SF, Ramage H, Cherry S, Hensley SE, Apostolidis SA, Huang AC, Vella LA, Unit UPCP, Betts MR, Meyer NJ, Wherry EJ (2020) Deep immune profiling of COVID-19 patients reveals distinct immunotypes with therapeutic implications. Science 369. doi: 10.1126/science.abc8511

Mehandru S, Merad M (2022) Pathological sequelae of long-haul COVID. Nat Immunol 23: 194–202. doi: 10.1038/s41590-021-01104-y

Merad M, Blish CA, Sallusto F, Iwasaki A (2022) The immunology and immunopathology of COVID-19. Science 375: 1122–1127. doi: 10.1126/science.abm8108

Nakanishi T, Farjoun Y, Willett J, Allen RJ, Guillen-Guio B, Zhou S, Richards JB (2022) Alternative splicing in the lung influences COVID-19 severity and respiratory diseases. medRxiv. doi: 10.1101/2022.10.18.22281202

Ong EZ, Chan YFZ, Leong WY, Lee NMY, Kalimuddin S, Haja Mohideen SM, Chan KS, Tan AT, Bertoletti A, Ooi EE, Low JGH (2020) A Dynamic Immune Response Shapes COVID-19 Progression. Cell Host Microbe 27: 879–882 e2. doi: 10.1016/j.chom.2020.03.021

Pairo-Castineira E, Clohisey S, Klaric L, Bretherick AD, Rawlik K, Pasko D, Walker S, Parkinson N, Fourman MH, Russell CD, Furniss J, Richmond A, Gountouna E, Wrobel N, Harrison D, Wang B, Wu Y, Meynert A, Griffiths F, Oosthuyzen W, Kousathanas A, Moutsianas L, Yang Z, Zhai R, Zheng C, Grimes G, Beale R, Millar J, Shih B, Keating S, Zechner M, Haley C, Porteous DJ, Hayward C, Yang J, Knight J, Summers C, Shankar-Hari M, Klenerman P, Turtle L, Ho A, Moore SC, Hinds C, Horby P, Nichol A, Maslove D, Ling L, McAuley D, Montgomery H, Walsh T, Pereira AC, Renieri A, Gen OI, Investigators IC, Initiative C-HG, and Me I, Investigators B, Gen CI, Shen X, Ponting CP, Fawkes A, Tenesa A, Caulfield M, Scott R, Rowan K, Murphy L, Openshaw PJM, Semple MG, Law A, Vitart V, Wilson JF, Baillie JK (2021) Genetic mechanisms of critical illness in COVID-19. Nature 591: 92–98. doi: 10.1038/s41586-020-03065-y

Pairo-Castineira E, Rawlik K, Klaric L, Kousathanas A, Richmond A, Millar J, Russell CD, Malinauskas T, Thwaites R, Stuckey A, Odhams CA, Walker S, Griffiths F, Oosthuyzen W, Morrice K, Keating S, Nichol A, Semple MG, Knight J, Shankar-Hari M, Summers C, Hinds C, Horby P, Ling L, McAuley D, Montgomery H, Openshaw PJM, Walsh T, Tenesa A, Scott RH, Caulfield MJ, Moutsianas L, Ponting CP, Wilson JF, Vitart V, Pereira AC, Luchessi A, Parra E, Cruz- Guerrero R, Carracedo A, Fawkes A, Murphy L, Rowan K, Law A, Hendry SC, Baillie JK (2022) GWAS and meta-analysis identifies multiple new genetic mechanisms underlying severe Covid-19. medRxiv. doi: 10.1101/2022.03.07.22271833

Pietzner M, Chua RL, Wheeler E, Jechow K, Willett JDS, Radbruch H, Trump S, Heidecker B, Zeberg H, Heppner FL, Eils R, Mall MA, Richards JB, Sander LE, Lehmann I, Lukassen S, Wareham NJ, Conrad C, Langenberg C (2022) ELF5 is a potential respiratory epithelial cell-specific risk gene for severe COVID-19. Nat Commun 13: 4484. doi: 10.1038/s41467-022-31999-6

Pruim RJ, Welch RP, Sanna S, Teslovich TM, Chines PS, Gliedt TP, Boehnke M, Abecasis GR, Willer CJ (2010) LocusZoom: regional visualization of genome-wide association scan results. Bioinformatics 26: 2336–7. doi: 10.1093/bioinformatics/btq419

Randolph HE, Fiege JK, Thielen BK, Mickelson CK, Shiratori M, Barroso-Batista J, Langlois RA, Barreiro LB (2021) Genetic ancestry effects on the response to viral infection are pervasive but cell type specific. Science 374: 1127–1133. doi: 10.1126/science.abg0928

Rocheleau G, Forrest IS, Duffy A, Bafna S, Dobbyn A, Verbanck M, Won HH, Jordan DM, Do R (2022) A tissue-level phenome-wide network map of colocalized genes and phenotypes in the UK Biobank. Commun Biol 5: 849. doi: 10.1038/s42003-022-03820-z

Schmiedel BJ, Rocha J, Gonzalez-Colin C, Bhattacharyya S, Madrigal A, Ottensmeier CH, Ay F, Chandra V, Vijayanand P (2021) COVID-19 genetic risk variants are associated with expression of multiple genes in diverse immune cell types. Nat Commun 12: 6760. doi: 10.1038/s41467-021-26888-3

Severe Covid GG, Ellinghaus D, Degenhardt F, Bujanda L, Buti M, Albillos A, Invernizzi P, Fernandez J, Prati D, Baselli G, Asselta R, Grimsrud MM, Milani C, Aziz F, Kassens J, May S, Wendorff M, Wienbrandt L, Uellendahl-Werth F, Zheng T, Yi X, de Pablo R, Chercoles AG, Palom A, Garcia-Fernandez AE, Rodriguez-Frias F, Zanella A, Bandera A, Protti A, Aghemo A, Lleo A, Biondi A, Caballero-Garralda A, Gori A, Tanck A, Carreras Nolla A, Latiano A, Fracanzani AL, Peschuck A, Julia A, Pesenti A, Voza A, Jimenez D, Mateos B, Nafria Jimenez B, Quereda C, Paccapelo C, Gassner C, Angelini C, Cea C, Solier A, Pestana D, Muniz-Diaz E, Sandoval E, Paraboschi EM, Navas E, Garcia Sanchez F, Ceriotti F, Martinelli-Boneschi F, Peyvandi F, Blasi F, Tellez L, Blanco-Grau A, Hemmrich-Stanisak G, Grasselli G, Costantino G, Cardamone G, Foti G, Aneli S, Kurihara H, ElAbd H, My I, Galvan-Femenia I, Martin J, Erdmann J, Ferrusquia-Acosta J, Garcia-Etxebarria K, Izquierdo-Sanchez L, Bettini LR, Sumoy L, Terranova L, Moreira L, Santoro L, Scudeller L, Mesonero F, Roade L, Ruhlemann MC, Schaefer M, Carrabba M, Riveiro-Barciela M, Figuera Basso ME, Valsecchi MG, Hernandez-Tejero M, Acosta-Herrera M, D’Angio M, Baldini M, Cazzaniga M, Schulzky M, Cecconi M, Wittig M, et al. (2020) Genomewide Association Study of Severe Covid-19 with Respiratory Failure. N Engl J Med 383: 1522–1534. doi: 10.1056/NEJMoa2020283

Sharif-Zak M, Abbasi-Jorjandi M, Asadikaram G, Ghoreshi ZA, Rezazadeh-Jabalbarzi M, Afsharipur A, Rashidinejad H, Khajepour F, Jafarzadeh A, Arefinia N, Kheyrkhah A, Abolhassani M (2022) CCR2 and DPP9 expression in the peripheral blood of COVID-19 patients: Influences of the disease severity and gender. Immunobiology 227: 152184. doi: 10.1016/j.imbio.2022.152184

Skrivankova VW, Richmond RC, Woolf BAR, Yarmolinsky J, Davies NM, Swanson SA, VanderWeele TJ, Higgins JPT, Timpson NJ, Dimou N, Langenberg C, Golub RM, Loder EW, Gallo V, Tybjaerg-Hansen A, Davey Smith G, Egger M, Richards JB (2021) Strengthening the Reporting of Observational Studies in Epidemiology Using Mendelian Randomization: The STROBE-MR Statement. JAMA 326: 1614–1621. doi: 10.1001/jama.2021.18236

Smith GD, Ebrahim S (2003) ’Mendelian randomization’: can genetic epidemiology contribute to understanding environmental determinants of disease? Int J Epidemiol 32: 1–22. doi: 10.1093/ije/dyg070

Soskic B, Cano-Gamez E, Smyth DJ, Ambridge K, Ke Z, Matte JC, Bossini-Castillo L, Kaplanis J, Ramirez-Navarro L, Lorenc A, Nakic N, Esparza-Gordillo J, Rowan W, Wille D, Tough DF, Bronson PG, Trynka G (2022) Immune disease risk variants regulate gene expression dynamics during CD4(+) T cell activation. Nat Genet. doi: 10.1038/s41588-022-01066-3

Tan LY, Komarasamy TV, Rmt Balasubramaniam V (2021) Hyperinflammatory Immune Response and COVID-19: A Double Edged Sword. Front Immunol 12: 742941. doi: 10.3389/fimmu.2021.742941

Wang Y, Guga S, Wu K, Khaw Z, Tzoumkas K, Tombleson P, Comeau ME, Langefeld CD, Cunninghame Graham DS, Morris DL, Vyse TJ (2022) COVID-19 and systemic lupus erythematosus genetics: A balance between autoimmune disease risk and protection against infection. PLoS Genet 18: e1010253. doi: 10.1371/journal.pgen.1010253

Wu L, Zhu J, Liu D, Sun Y, Wu C (2021) An integrative multiomics analysis identifies putative causal genes for COVID-19 severity. Genet Med 23: 2076–2086. doi: 10.1038/s41436-021-01243-5

Yoshiji S, Butler-Laporte G, Lu T, Willett JDS, Su CY, Nakanishi T, Morrison DR, Chen Y, Liang K, Hultstrom M, Ilboudo Y, Afrasiabi Z, Lan S, Duggan N, DeLuca C, Vaezi M, Tselios C, Xue X, Bouab M, Shi F, Laurent L, Munter HM, Afilalo M, Afilalo J, Mooser V, Timpson NJ, Zeberg H, Zhou S, Forgetta V, Farjoun Y, Richards JB (2023) Proteome-wide Mendelian randomization implicates nephronectin as an actionable mediator of the effect of obesity on COVID-19 severity. Nat Metab 5: 248–264. doi: 10.1038/s42255-023-00742-w

Zheng J, Haberland V, Baird D, Walker V, Haycock PC, Hurle MR, Gutteridge A, Erola P, Liu Y, Luo S, Robinson J, Richardson TG, Staley JR, Elsworth B, Burgess S, Sun BB, Danesh J, Runz H, Maranville JC, Martin HM, Yarmolinsky J, Laurin C, Holmes MV, Liu JZ, Estrada K, Santos R, McCarthy L, Waterworth D, Nelson MR, Smith GD, Butterworth AS, Hemani G, Scott RA, Gaunt TR (2020) Phenome-wide Mendelian randomization mapping the influence of the plasma proteome on complex diseases. Nat Genet 52: 1122–1131. doi: 10.1038/s41588-020-0682-6

Zhou S, Butler-Laporte G, Nakanishi T, Morrison DR, Afilalo J, Afilalo M, Laurent L, Pietzner M, Kerrison N, Zhao K, Brunet-Ratnasingham E, Henry D, Kimchi N, Afrasiabi Z, Rezk N, Bouab M, Petitjean L, Guzman C, Xue X, Tselios C, Vulesevic B, Adeleye O, Abdullah T, Almamlouk N, Chen Y, Chasse M, Durand M, Paterson C, Normark J, Frithiof R, Lipcsey M, Hultstrom M, Greenwood CMT, Zeberg H, Langenberg C, Thysell E, Pollak M, Mooser V, Forgetta V, Kaufmann DE, Richards JB (2021) A Neanderthal OAS1 isoform protects individuals of European ancestry against COVID-19 susceptibility and severity. Nat Med 27: 659–667. doi: 10.1038/s41591-021-01281-1

